# The Impact of Digital Adherence Technologies on Health Outcomes in Tuberculosis: A Systematic Review and Meta-Analysis

**DOI:** 10.1101/2024.01.31.24302115

**Authors:** Mona S. Mohamed, Miranda Zary, Cedric Kafie, Chimweta I. Chilala, Shruti Bahukudumbi, Nicola Foster, Genevieve Gore, Katherine Fielding, Ramnath Subbaraman, Kevin Schwartzman

## Abstract

**Background:** Suboptimal tuberculosis (TB) treatment adherence may lead to unsuccessful treatment and relapse. Digital adherence technologies (DATs) may allow more person-centric approaches for supporting treatment. We conducted a systematic review (PROSPERO-CRD42022313166) to evaluate the impact of DATs on health outcomes in TB.

**Methods:** We searched MEDLINE, Embase, CENTRAL, CINAHL, Web of Science and preprints from medRxiv, Europe PMC, and clinicaltrials.gov for relevant literature from January 2000 to April 2023. We considered experimental or cohort studies reporting quantitative comparisons of clinical outcomes between a DAT and the standard of care in each setting.

**Results:** Seventy studies (total 58,950 participants) met inclusion criteria. They reported SMS-based interventions (k=18 studies), feature phone-based interventions (k=7), medication sleeves with phone calls (branded as “99DOTS,” k=5), video-observed therapy (VOT; k=17), smartphone-based interventions (k=5), digital pillboxes (k=18), ingestible sensors (k=1), and interventions combining 2 DATs (k=1). Overall, the use of DATs was associated with more frequent treatment success in TB disease (OR = 1.18 [1.06, 1.33]; I^2^ = 66%, k = 46), a decrease in loss to follow up (OR = 0.71 [0.53, 0.94]; I^2^ = 80%, k = 36) and an increase in adverse event reporting (OR = 1.53 [1.26, 1.86]; I^2^ = 0%, k = 9). VOT was associated with an increased likelihood of treatment success in TB disease (OR 1.54 [1.09; 2.19]; I^2^ = 0%, k = 8) and treatment completion in TB infection (OR 4.69 [2.08; 10.55]; I^2^ = 0%, k = 2) as well as an increased frequency of adverse event reporting (OR = 1.79 [1.27; 2.52]; I^2^ = 34%, k = 4). Other interventions involving smartphone technologies were associated with increased treatment success in TB disease (OR 1.98 [1.07; 3.65]; I^2^ =56%, k = 5) and a decreased frequency of loss to follow up (OR = 0.31 [0.13; 0.77]; I^2^ = 36%, k = 5). Digital pillboxes were also associated with an improvement in treatment success (OR = 1.32 [1.00; 1.73]; I^2^ = 71%, k = 11). However, improved treatment success was only seen in high- and upper middle-income countries. SMS-based interventions, feature-phone interventions and 99DOTS were not associated with improvements in short-term clinical outcomes.

**Conclusion:** Certain DATs--notably VOT and smartphone-based interventions, in higher income settings and sometimes combined with other supportive measures—may be associated with improvements in treatment success and losses to follow-up, compared to standard care. However, evidence remains highly variable, and generalizability limited. Higher quality data are needed.

## Introduction

Tuberculosis (TB) is currently the second leading infectious cause of death, eclipsed only by the COVID-19 pandemic. About one-quarter of the world’s population is estimated to have been infected with *Mycobacterium tuberculosis*.[1] In 2022, an estimated 10.6 million people developed TB disease worldwide, of whom 1.3 million died.[2]

Suboptimal adherence to treatment for TB disease may lead to treatment failure, increased risk of death or relapse. To improve adherence, the World Health Organization in 1994 endorsed directly observed therapy (DOT) where health workers, community volunteers or family members observe and record patients taking each dose.[3] Digital adherence technologies (DATs) may facilitate more patient-centric, community rather than facility-based, and less intrusive approaches for monitoring TB medication adherence than existing DOT models.[4] Depending on the setting and care model, some studies have suggested that DAT-based treatment may be more effective than standard care in achieving favorable treatment outcomes.[5] However, findings regarding health outcomes have been inconsistent and may vary by country income level and technology type. [6–8]

We conducted a systematic review and meta-analysis to evaluate the impact of DATs on health outcomes in persons treated for TB disease and TB infection.

## Methods

Our systematic review and meta-analysis was conducted and reported using the Preferred Reporting Items for Systematic Reviews and Meta-Analyses (PRISMA) guidelines[9] (S1 checklist), and followed a protocol registered as **PROSPERO-CRD42022313166.**

### Search Strategy and study selection

On April 25^th^, 2023 (updated from April 14^th^, 2022), we searched MEDLINE(Ovid), Embase (Ovid), CENTRAL (Wiley), CINAHL, Web of Science Core Collection, Europe PMC preprints (including MedRxiv) and ClinicalTrials.gov (for unpublished clinical trials). from January 1^st^, 2000, to April 14^th^, 2023. The search was conducted by a health sciences librarian (GG) to ensure that the most suitable search terms were used. Key elements included terms that denoted **tuberculosis** (disease or infection); **digital technologies** and tools (including mobile phone, smartphone, video observation, medication monitors, text messaging, etc.) and **health outcomes** (microbiologic conversion, treatment success, microbiologic cure, treatment failure, death, loss to follow-up, relapse/ recurrence, subsequent TB disease after treatment for latent TB infection, medication adherence, and patient-reported outcomes). More detailed information about the search strategy and results is provided in S1 Table.

We also hand-searched conference abstracts from the Union World Conference on Lung Health from 2004 to 2022 and cross-checked reference lists and citations of included studies as a supplementary approach to identify further data sources.

We followed a PICOS format (participants, interventions, comparator, outcomes, and study design) to formulate the clinical question used for including studies in the systematic review.

#### Study design

We included randomized controlled trials (RCTs), quasi-experimental studies, and cohort studies.

#### Population

The population of interest were individuals diagnosed and treated for TB disease or infection, including persons at risk of unfavorable outcomes (e.g., those with drug-resistant disease, those living with HIV, children).

#### Interventions

We considered studies that addressed DATs intended to monitor and support TB treatment adherence, including medication dosing but also visit attendance and continued follow-up. This included reminding patients to take their medications, facilitating digital observation of pill-taking, compiling patient dosing histories and identifying adherence patterns, which facilitate patient-centric approaches for monitoring TB medication adherence. We included all such interventions whether they engaged persons treated for TB, providers, or both. We excluded studies where the technology was an electronic health record used only to log visit attendance or if the intervention consisted only of non-automated “routine telephone calls” to patients. A detailed definition for eligible DAT interventions was developed before study selection and is provided in S2 Table.

#### Comparator

To be eligible for inclusion, studies had to report a comparator arm involving standard care in the same settings. This included various forms of directly observed therapy (DOT), self-administered therapy (SAT) or a combination of the two. Approaches to DOT used as the standard of care in different studies varied substantially and could include facility-based DOT (where people with TB had to visit clinics or other health facilities for in-person observation of dosing), field-based DOT (where a healthcare provider or other treatment supporter visited people with TB at their home or another location for observation of dosing), or family DOT (where a family member or other local supporter observed dosing).

#### Outcomes

We considered short- and long-term clinical outcomes, medication adherence, and patient-reported outcomes. Short-term clinical outcomes were treated as binary (yes/no) and included: treatment success, treatment completion, cure, loss to follow up, treatment failure, death during treatment, adverse events, emergence of anti-tuberculous drug resistance, microbiological conversion of sputum smear, completion of intensive phase treatment and composite outcomes of favorable or unfavorable outcomes. Short-term clinical outcomes are defined in S3 Table.

Long-term clinical outcomes were also treated as binary, and included recurrence or relapse (TB disease), defined as a repeat diagnosis of TB disease within 2 years of successful treatment; post-treatment death, defined as death within 2 years of successful treatment for TB disease; and subsequent risk of TB disease, defined as development of TB disease within 5 years of treatment for TB infection.

Adherence was considered as a binary, ordinal, or continuous outcome depending on the measures used in specific studies: These included indirect measures of medication adherence such as pharmacy refill visits; or more direct measures of medication ingestion as indicated by persons with TB themselves, DAT and DOT records, or biologic testing (e.g. drug metabolites in urine). Patient-reported outcomes were based on suitably validated instruments, administered to persons using or not using DATs, at comparable time points during/after treatment. They included: health-related quality of life (HRQoL), functional status, perceived stigma and satisfaction.

To be included, studies had to report quantitative comparisons of clinical outcomes between a DAT and standard care with at least 20 participants using the DAT. Clinical outcomes could include treatment outcomes, other health outcomes and/or patient reported outcomes. We included data from full-text publications, preprints, or abstracts without any language restriction. We excluded review articles and studies that reported purely qualitative information.

After deduplication of items initially retrieved from our search, all remaining items were imported into Rayyan software (Rayyan, Cambridge USA). Selection of studies involved two levels of screening conducted in parallel by two independent reviewers (among MM, MZ, CK, CC, SB, NF). In the first-level screen, all titles and abstracts were screened: all studies addressing TB and DATs were included. A third reviewer (KS, KF, or RS) resolved conflicts. In the second-level screen, two independent reviewers (MM, MZ, CK, SB and NF) screened the full text of studies retained from the title and abstract review. Articles were excluded if they did not meet the inclusion criteria. In the event of discordant judgment regarding suitability, a third reviewer (KS) resolved conflicts. Excluded full-texts were stored and the reasons for their exclusion were documented.

### Data Extraction

Data extraction was conducted independently by two reviewers (among MM, MZ, CK, CC) into a standardised template. Disagreements between reviewers were resolved by consensus and discussion with a third reviewer (KS) when needed. Data extracted included study characteristics (design, year of study, country, setting, sample size, inclusion and exclusion criteria, limitations, strengths and conclusion), population characteristics (age, sex, number of participants in each group, TB condition, drug resistance, HIV co-infection), DAT intervention and comparator (type, duration, frequency of the intervention, approach if suboptimal adherence was detected), and outcomes (number of events in each group, measures of effect (means ratios, risk ratios, or odds ratios with respective confidence intervals).

### Quality Assessment

Each eligible study was evaluated by two reviewers (MM, MZ, CK, CC) independently to assess methodological quality. We judged trials to be either at a low, high, or unclear risk of bias in seven categories using Cochrane’s Assessment of Risk of Bias. (S4 Table)

Similarly, two reviewers independently evaluated each cohort study using the Newcastle Ottawa Scale. (S5 Table) Disagreements were resolved through discussion between the two reviewers, and with a third reviewer (KS) when needed.

### Data Synthesis

For each included study, data were summarized in tabular format to highlight study and participant characteristics, as well as key features of the DAT and comparator interventions reported. Results for short-term clinical outcomes were pooled and summarized quantitatively. When treatment success (which includes both successful completion and microbiologic cure) and treatment completion were reported in the same study, we focused on treatment success for purposes of meta-analysis. When only treatment completion was reported in a study, we used those data when pooling the results for treatment success.

Results were pooled using an inverse-variance weighted random effects model to estimate overall odds ratios and 95% confidence intervals. For short-term clinical outcomes, the odds ratio in individual studies was calculated from the number of events in the intervention and control groups. When the direct estimate required for the analysis – an odds ratio with its 95% confidence intervals-was provided by the authors, it was used when pooling the data. Risk ratios in individual studies were used when odds ratios were not reported. The reciprocal of odds ratio for unfavorable outcomes was used to calculate the odds ratio for treatment success in studies that addressed unfavorable outcomes only, and did not report treatment success. For cluster-randomized trials and stepped-wedge trials we used the direct estimate from analyses that properly accounted for the cluster design. When an effect measure was not reported, available crude data were used to estimate odds ratios and 95% confidence intervals.

Between-study variance was estimated by the DerSimonian-Laird method [10]. The meta-analysis was conduced using R version 4.2.3, R Foundation for Statistical Computing, Vienna, Austria, using the *metafor*[11] and *meta*[12] packages. When substantial heterogeneity was present (I^2^ > 50%) we conducted subgroup analyses to explore potential sources.

Pre-specified subgroup analyses considered the specific DAT interventions as well as income levels in the countries where studies were conducted. Country income level was classified as high, upper-middle, lower-middle or low according to the classification of the World Bank.[13] When the number of pooled studies exceeded 10, publication bias was assessed qualitatively by visual inspection of funnel plot symmetry and quantitatively using Egger’s linear regression.[14] A sensitivity analysis was completed with and without data extracted from conference abstracts to confirm the robustness of the results.

Treatment adherence was summarized in tabular form and described narratively. Odds ratios for adherence results with sufficiently homogeneous assessment measures were presented in forest plots without pooling. When adherence was reported in binary terms, odds ratios or risk ratios provided by the authors were used, or the odds ratio was estimated from the number of events in the intervention and comparison groups. For continuous adherence measurements, the standardized mean difference was calculated and transformed to the log odds using the Hasselblad and Hedges method.[15] As reports were much sparser, results for long-term clinical outcomes, and patient reported outcomes were summarized in tabular form and/ or described narratively following PRISMA guidelines for best practices.

### Grading of evidence

Finally, we rated the quality of evidence for each outcome we meta-analyzed, based on the Grading of Recommendations, Assessment, Development and Evaluations (GRADE) approach which considers study limitations, inconsistency of results, indirectness of evidence, imprecision, and publication bias.

## Results

Our search yielded 9,964 records as shown in Figure 1. After removing 2,580 duplicate records, a further 6,698 items were excluded after title and abstract review, as they did not address TB and/or DATs; 683 full texts were therefore retained for further eligibility screening, of which 56 studies met inclusion criteria. Our search of the Union Conference abstracts identified 13 additional studies, and one further study was identified after reviewing citations of all included studies, for a total of 70 included studies. These 70 studies reported on 67 unique study cohorts.[16–18]

**Fig 1.**
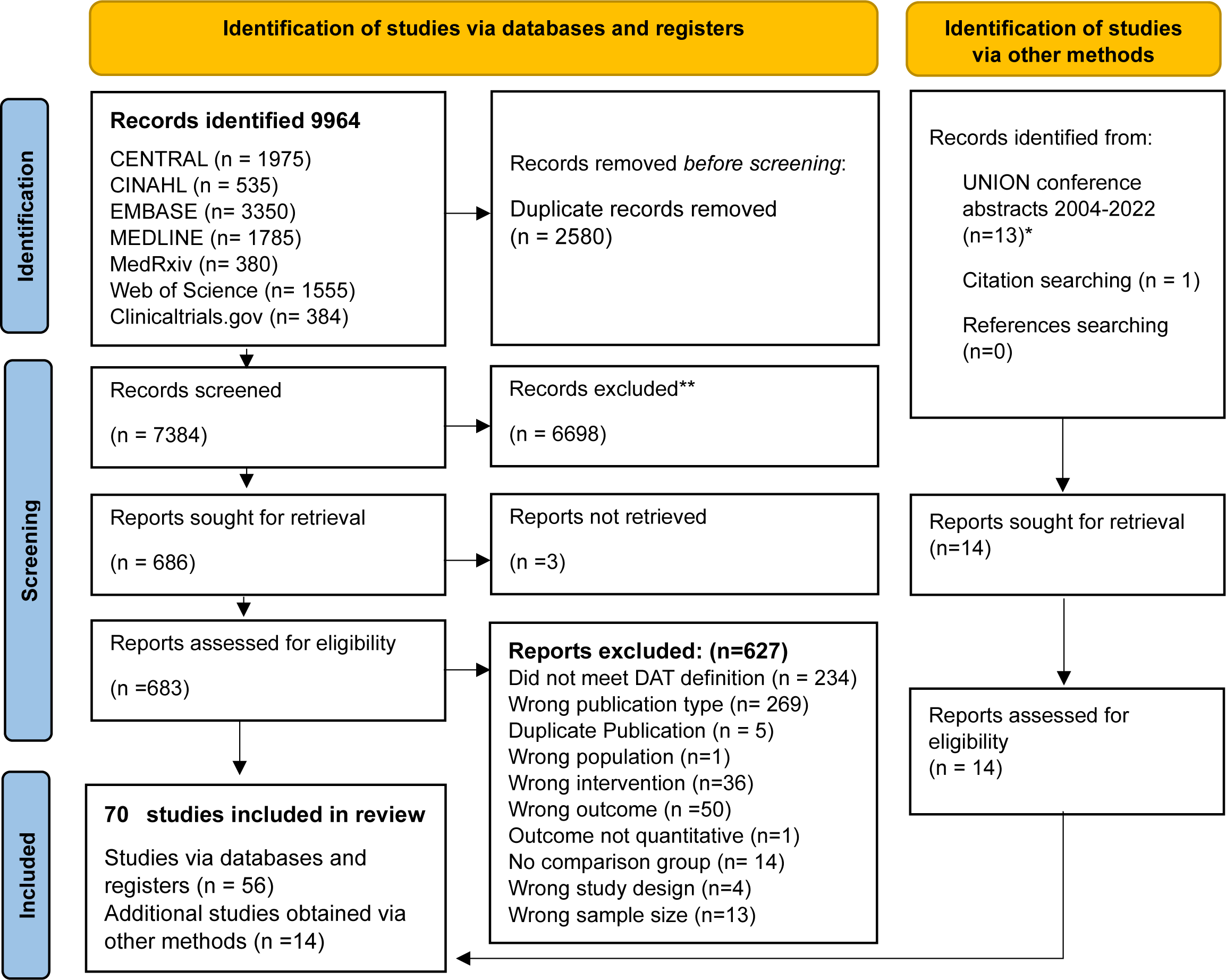
The PRISMA flow diagram explaining the flow of information through the different phases of the systematic review. *One UNION conference abstract was published in March 2023 as a Lancet preprint that was not caught in the search via Europe PMC database. The preprint was included instead.

### Characteristics of the included studies

Of the 70 included studies, 38 were randomized controlled trials (RCTs) (including one cross-over trial), five were quasi-experimental studies, and 27 were cohort studies. In those studies, a total of 29,228 participants used DATs while 29,722 others received standard care. Reports addressed eight types of DAT intervention: SMS-based interventions (k=18), feature phone-based interventions (k=7), medication sleeves with phone calls (hereafter referred to by its brand name “99DOTS” (k=5), video-observed therapy VOT (synchronous and/or asynchronous) (k=17), other interventions based on smartphone technology (k=5), digital pillboxes (medication event reminder monitor (MERM)) (k=18), ingestible sensors (k=1) and interventions involving a combination of DATs (k=1).

VOT and ingestible sensors were exclusively used in high income countries (HICs) and upper-middle income countries (UMICs), while DATs used in low-income countries (LICs) included SMS, feature phone-based interventions, digital pillboxes and 99DOTS.

Of the 70 included studies, 9 used facility-based DOT as the comparator, three gave the patients the choice between facility- or field-based DOT, 9 used field-based DOT provided by a health worker, of which three allowed observation at home, two used DOT provided by a family member or friend, 15 used DOT but did not provide further details, 9 used self-administered treatment, 8 used a combination, and 15 were unclear.(S6 Table)

Among participants in the included studies, 99DOTS was provided to the largest number (42.5%). A summary of study and participant characteristics is provided in Table 1, while Fig 2 highlights DATs reported according to country and income level. Detailed characteristics of each study, participants, interventions used, and outcomes reported are listed in (S6-S8 Tables).

**Fig 2.**
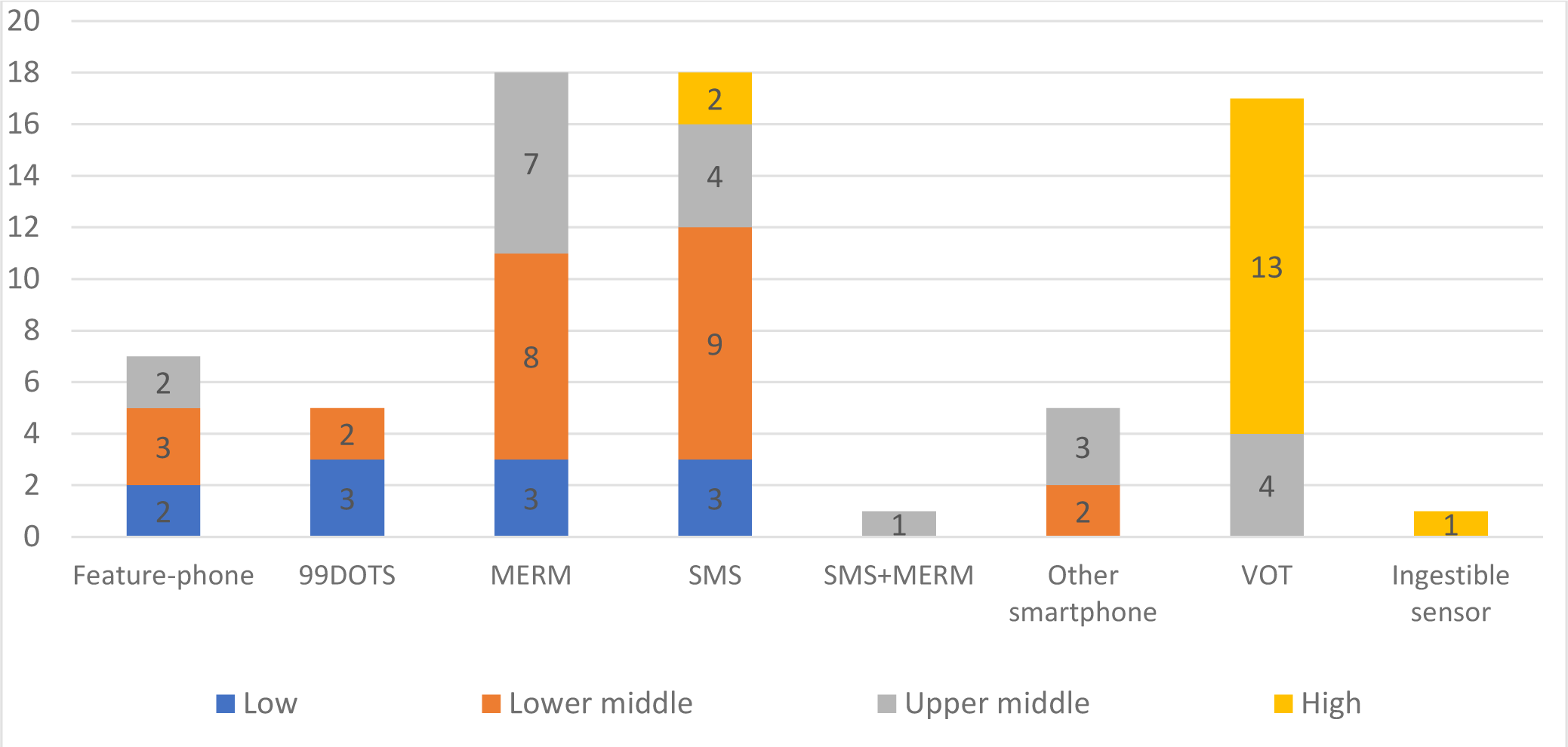
Distribution of DATs by country-income-level. **MERM** digital pillboxes, **SMS+ MERM** a combination of SMS-based intervention and digital pillboxes, **VOT** video observed therapy. One study included 3 different DAT intervention arms, and one was conducted in several upper-middle and high-income countries.

**Table 1.** Summary of characteristics of included studies and participants.

| Subgroups | Number of studies | Number of participants using DAT | Number of participants in SOC arm |
| --- | --- | --- | --- |
| **All studies included** | **70** | **29,228** | **29,722** |
| **Publication type** |  |  |  |
| Full-text paper | 54 | 14,801 | 14,027 |
| Preprint | 2 | 8,557 | 7,930 |
| Abstract | 13 | 5,301 | 7,230 |
| Research letter | 1 | 569 | 535 |
| **Study design** |  |  |  |
| RCT | 38 | 12,422 | 10,453 |
| Quasi-experimental | 5 | 664 | 783 |
| Cohort study* | 27 | 16,142 | 18,486 |
| **DAT type** |  |  |  |
| SMS | 18** | 4,515 | 4,788** |
| VOT | 17 | 1,628 | 2,502 |
| MERM | 18** | 6,457 | 8,884** |
| Feature-phone | 7 | 1,760 | 1,574 |
| 99DOTS | 5 | 12,386 | 11,284 |
| Other smartphone | 5 | 1,377 | 1,774 |
| Ingestible sensor | 1 | 41 | 20 |
| SMS+MERM | 1** | 1,064 | 1,104** |
| **Country-income level** |  |  |  |
| Low | 11 | 4,113 | 3,681 |
| Lower-Middle | 24 | 17,062 | 19,394 |
| Upper-Middle | 19 | 6,492 | 4,531 |
| Upper-Middle and High | 1 | 328 | 337 |
| High | 15 | 1,233 | 1,779 |
| **TB disease vs. infection** |  |  |  |
| TB disease | 65 | 28,553 | 28,688 |
| TB infection | 4 | 628 | 987 |
| Both | 1 | 47 | 47 |
| **TB resistance for TB disease** |  |  |  |
| Drug-susceptible | 40 | 22,797 | 24,172 |
| Multi-drug resistant | 2 | 151 | 211 |
| Both drug-sensitive and drug-resistant | 2 | 138 | 79 |
| Unknown | 21 | 5,467 | 4,226 |

\* One study was a comparative cross-sectional study with retrospective data
\*\* Liu et al 2015 included 3 DAT arms (SMS only, MERM only and SMS and MERM (combined) and 1 control arm

### Risk of bias in included studies

Performance bias was common in most RCTs, as it was not feasible to mask participants to the interventions used. High risk of detection bias was also present in over 30% of the studies, as outcome assessors were not blinded. All studies had either low or unclear risk of selection bias. (S1 and S2 Figs).

More than 60% of the included observational studies showed high risk of bias related to participant representativeness. Many studies required participants to be literate, have access to a mobile phone, demonstrate digital literacy, and/or have a reliable internet connection in order to participate. (S3 Fig, S9 and S10 Tables)

### Short-term clinical outcomes in TB disease

A summary of findings for the effect of digital technologies on short-term clinical outcomes in TB disease is provided in Figures 3 and 4 and S17 Table.

**Fig 3.**
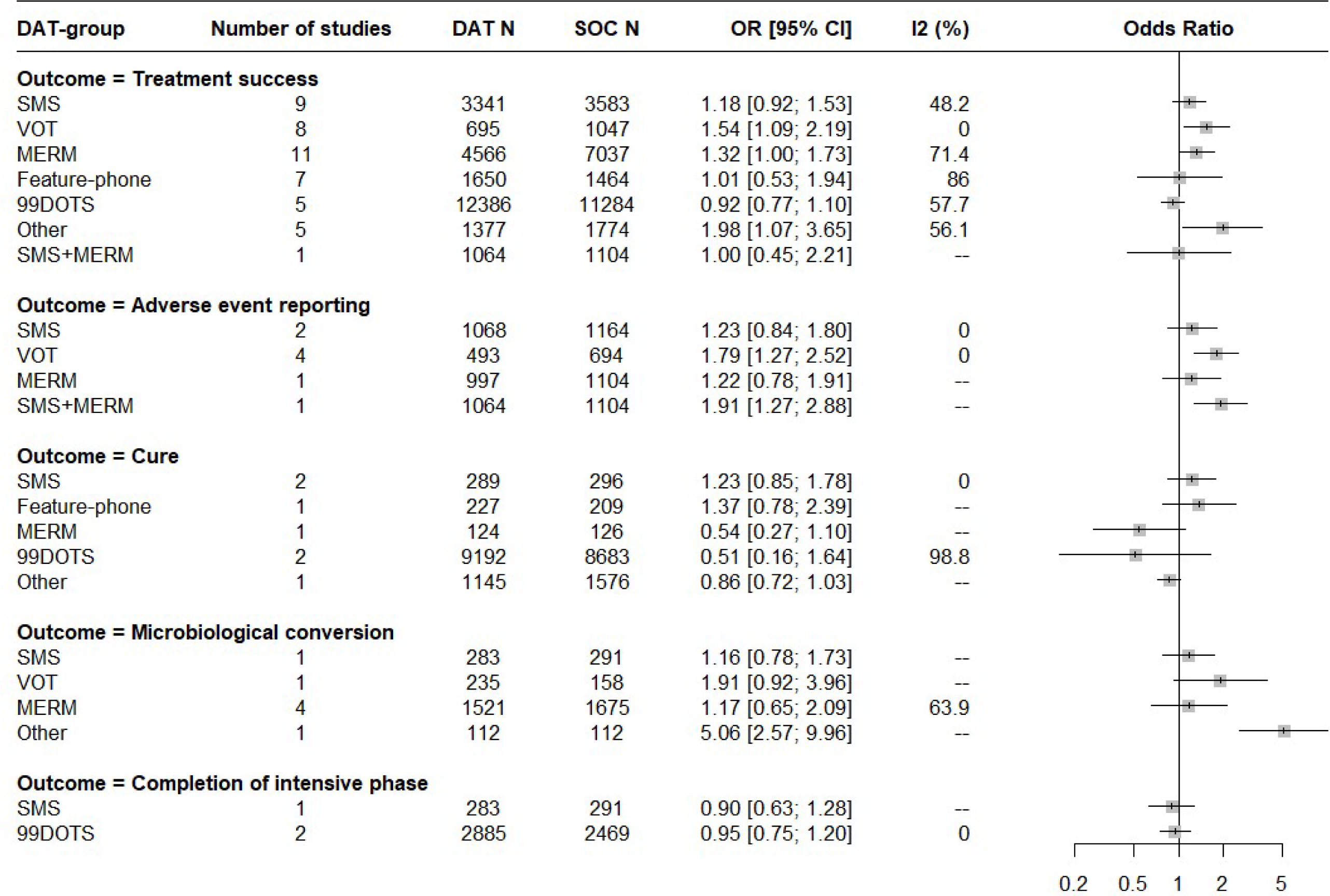
Pooled estimates for favorable short-term clinical outcomes in TB disease stratified by type of DAT.

**Fig 4.**
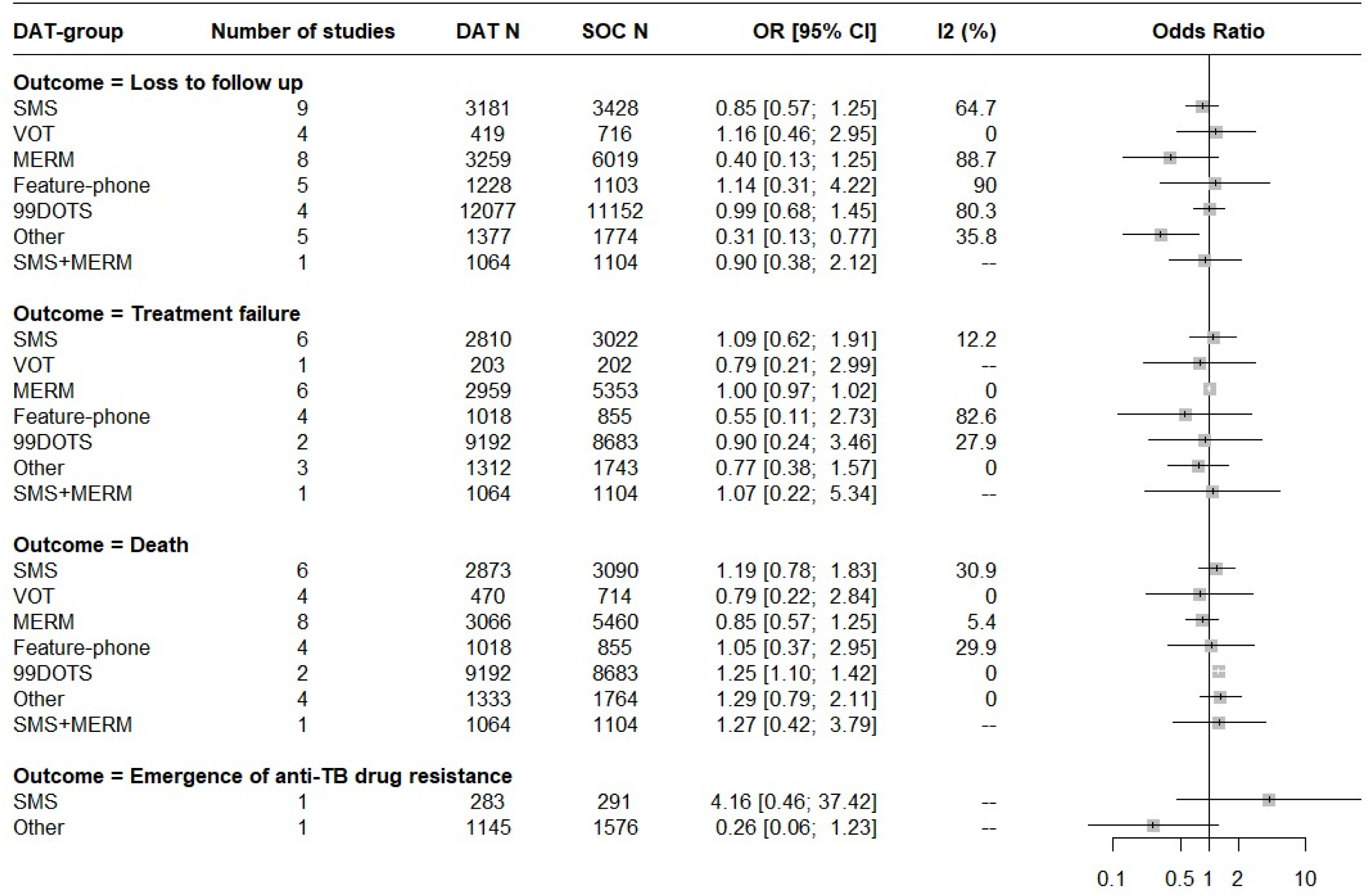
Pooled estimates for unfavorable short-term clinical outcomes in TB disease stratified by type of DAT.

#### Treatment success

Overall, the use of digital technologies was associated with a small increase in treatment success, with substantial heterogeneity across studies (OR = 1.18 [1.06, 1.33]; I^2^ = 66%, k = 46, [very low certainty evidence]). Video-observed therapy (OR 1.54 [1.09; 2.19]; I^2^ = 0%, k = 8, [low certainty evidence]) and smartphone-based technologies (other) (OR 1.98 [1.07; 3.65]; I^2^ =56%, k = 5, [low certainty evidence]) were each associated with an increased likelihood of treatment success for persons with TB disease, as were digital pillboxes) (OR 1.32 [1.00; 1.73]; I^2^ =86%, k = 11, [low certainty evidence]) while other DAT types were not. Further subgroup analysis suggested that VOT (OR = 1.55 [1.04; 2.31]; I^2^ = 0%, k = 5) were associated with greater treatment success in high-income countries, while smartphone-based technologies (OR = 2.81 [1.11; 7.14]; I^2^ = 33%, k = 3) were associated with increased treatment success in upper-middle income countries. (S11 and S12 Tables, S4 Fig) A supplementary meta-analysis showed that DATs used in high-income countries (OR = 1.55 [1.04; 2.31]; I^2^ = 0%, k = 5) or upper-middle income countries (OR = 1.23 [1.02; 1.50]; I^2^ = 36%, k = 18) were associated with improvement in treatment success. (S16 Fig)

#### Loss to follow up

Similarly, DATs overall were associated with a significant decrease in the proportion of patients lost to follow up, again with substantial heterogeneity (OR = 0.71 [0.53, 0.94; I^2^ = 80%, k = 36, [low certainty evidence]). Among the specific DATs, smartphone-based interventions were associated with decreased likelihood of loss to follow up (OR = 0.31 [0.13; 0.77]; I^2^ = 36%, k = 5, [moderate certainty evidence]). (S13 Table, S5 Fig) In low-income countries, loss to follow up was less frequent among persons using any DAT than among those who received the local standard of care (S17 Fig).

#### Treatment failure and death

None of the digital technology interventions was associated with reductions in treatment failure. (S14 Table, S8 Fig). However, digital technologies overall (OR= 1.19 [1.07, 1.32]; I^2^=0%, k=29, [low certainty evidence]) and medication sleeves with phone calls (99DOTS) (OR = 1.25 [1.10, 1.42; I^2^ = 0%, k = 2, [low certainty evidence]) were associated with a small increase in the likelihood of death. (S15 Table, S7 Fig) Importantly, clustering of participants could not be accounted for in this specific analysis.

#### Adverse events

The use of digital technologies was associated with an increase in the number of persons reporting adverse events (OR = 1.53 [1.26; 1.86]; I^2^ = 0%, k = 8, [moderate certainty evidence]). Subgroup analysis showed that VOT (OR = 1.79 [1.27; 2.52]; I^2^ = 0%, k = 4, [moderate certainty evidence]) was associated with an increased frequency of adverse event reporting. The combination of SMS and MERM (OR = 1.91 [1.27; 2.88]; I^2^ = N/A, k = 1) also significantly increased the number of adverse events reported. (S16 Table, S8 Fig). While in most studies the number of adverse events corresponded to the number of persons who reported any adverse event[19–22], in one study it referred specifically to persons for whom adverse events led to longer TB treatment duration [23], and in another study it referred to persons for whom adverse events led to treatment interruption.[24] Abdominal pain, nausea, and vomiting were the most common adverse events reported. [19,20]

#### Cure

Seven studies reported on cure; none of the digital technology interventions was associated with a difference in microbiologic cure among persons with TB disease. (S9 Fig)

#### Microbiologic conversion

Nine studies assessed microbiologic conversion of sputum smears or cultures in persons with TB disease. Overall, there was no significant difference between the DAT intervention and comparison groups. While eight studies addressed the proportion of patients with microbiologic conversion in the DAT group compared to standard care, one study [25] reported the mean time to culture conversion in the VOT group (47 days) vs. the standard of care group (48 days). (S17 Table, S10 Fig)

#### Completion of intensive phase treatment

Three studies reported on completion of intensive phase treatment for TB disease. None showed a significant association with DAT use. (S11 Fig)

#### Emergence of antibiotic resistance

Two studies reported on emergence of anti-TB antibiotic resistance during treatment. Neither showed a significant association with DAT use. (S12 Fig)

### Publication bias

Publication bias was assessed using funnel plots for treatment success, loss to follow up, treatment failure and death. Egger’s test indicated asymmetry for treatment success (Egger’s β_0_ = 0.66 [0.16 to 1.16]; t_(44)_ = 2.58; p = 0.01) suggesting potential publication bias reflected by the relative paucity of larger studies with negative findings. (Fig 6) No publication bias was detected for loss to follow-up, treatment failure or death. (S13-15 Figs)

### Sensitivity analysis

A sensitivity analysis was conducted after excluding studies reported as conference abstracts. Overall findings were mostly similar, except that there was no longer a significant association between DAT use in general and loss to follow-up. Similarly, there was no longer a significant association between the smartphone technology-based interventions and treatment success.

### Short-term clinical outcomes in TB infection

#### Treatment completion

Four studies (1615 participants) addressed the use of DATs (2 SMS[26,27], 2 VOT[28,29]) in TB infection. Three studies were conducted in HIC and one study included outpatient tuberculosis clinics in three HIC and one UMIC[27]. The definition of treatment completion varied across the studies. Some studies involving weekly isoniazid-rifapentine treatment defined it as ingestion of 11 doses within 16 weeks[27,28], while another study required ingestion of at least 80% or 90% of planned medication doses within specified time intervals, i.e. ⩾80% or 90% of prescribed INH doses within 12 months, or ⩾80% or 90% of prescribed rifampin doses within 6 months.[26] Overall, DAT use was associated with a significant increase in treatment completion. (OR = 1.77 [0.85, 3.66; I^2^ = 77%, k = 4, [low certainty evidence]). (S18 Table, Fig 5). A subgroup analysis showed that only VOT was associated with improved treatment completion. (OR = 4.69 [2.08, 10.55; I^2^ = 0%, k = 2)

**Fig 5.**
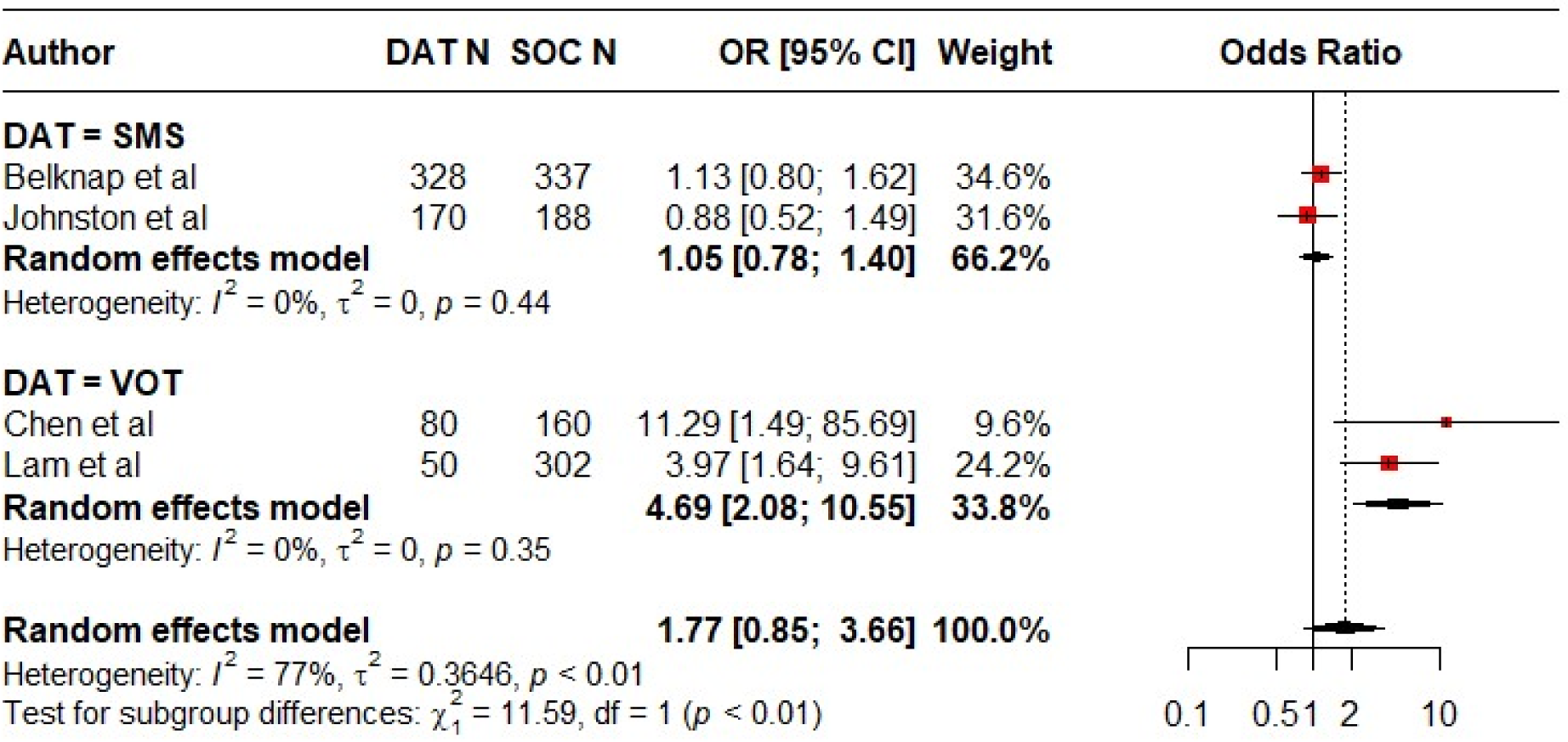
Forest plot showing treatment completion in DAT groups compared to standard of care among persons with TB infection.

**Fig 6.**
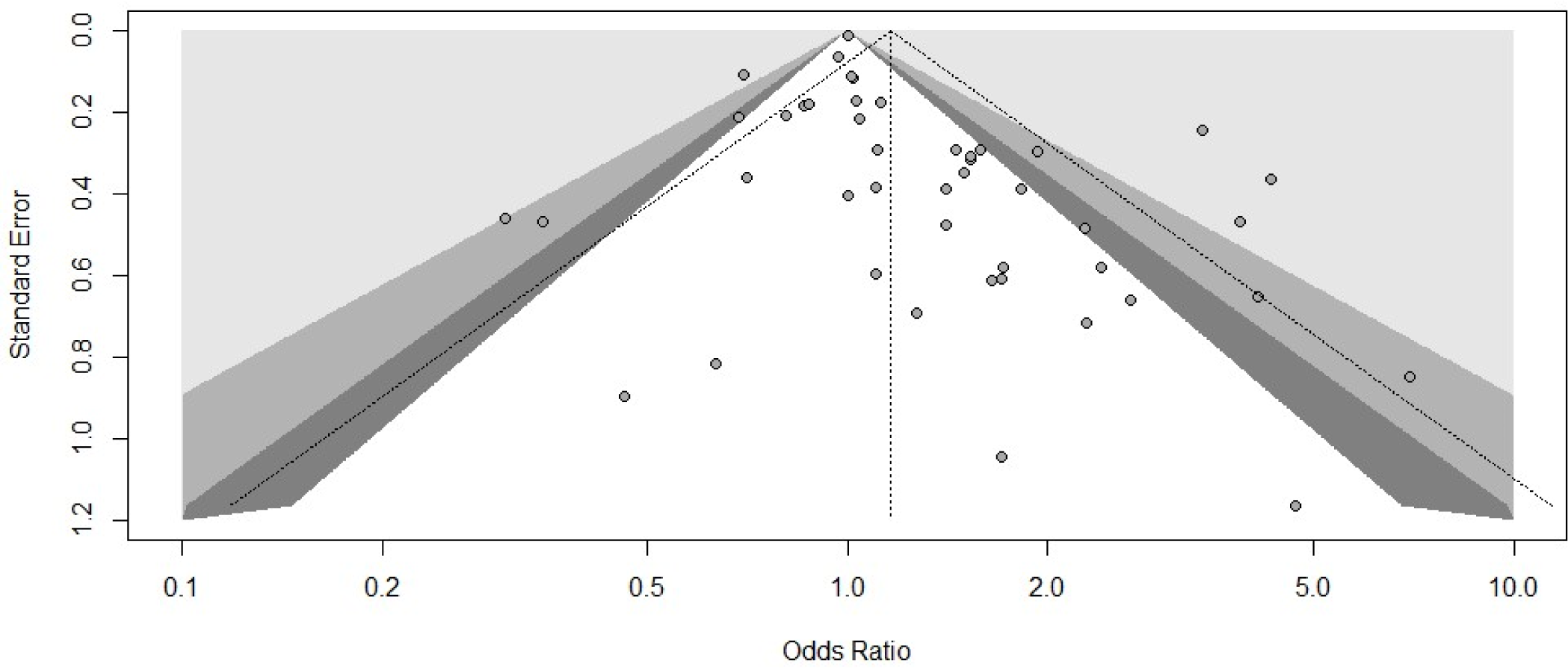
Funnel plot of included studies for the assessment of publication bias using Egger’s test. It shows studies comparing treatment success in patients using DATs versus standard care. Each point represents an individual study. Varying levels of statistical significance of the studies are indicated by the shading; the unshaded region in the middle corresponds to p-values greater than 0.10, the dark gray-shaded region corresponds to p-values between 0.05 and 0.10, the medium gray-shaded region corresponds to p-values between 0.01 and 0.05, and the region outside of the funnel corresponds to p-values below 0.01.

#### Loss to follow up and adverse events

Results for loss to follow-up and adverse event reporting were inconsistent: one study suggested that VOT was associated with an increase in loss to follow-up[29], while another suggested that a two-way SMS-based intervention increased the number of adverse events reported.[26]

### Long-term clinical outcomes

Two cohort studies [22,30]and one RCT [31]examined the frequency of recurrence after treatment for TB disease. Of these, only one cohort study suggested a lower rate of recurrence in the DAT group. The others found no difference.[30]

None of the included studies assessed death after treatment completion, or subsequent risk of TB disease among persons treated for TB infection.

### Adherence

There was substantial methodological heterogeneity among studies reporting on adherence. Key differences included the measurements used to assess adherence (S20 Fig), the interval over which adherence was measured as well as the timing of the assessment. (S19 Table)

Of 19 studies examining SMS interventions, 13 (68%) reported adherence measures. In eight, this reflected patient self-reports. Three studies assessed adherence in terms of (timely) clinic attendance and one used pill-counts. When compared to simultaneous urine tests for isoniazid metabolites self-reports over-reported medication ingestion. [32] Association of the SMS interventions with treatment adherence was inconsistent. (Fig 7)

**Fig 7.**
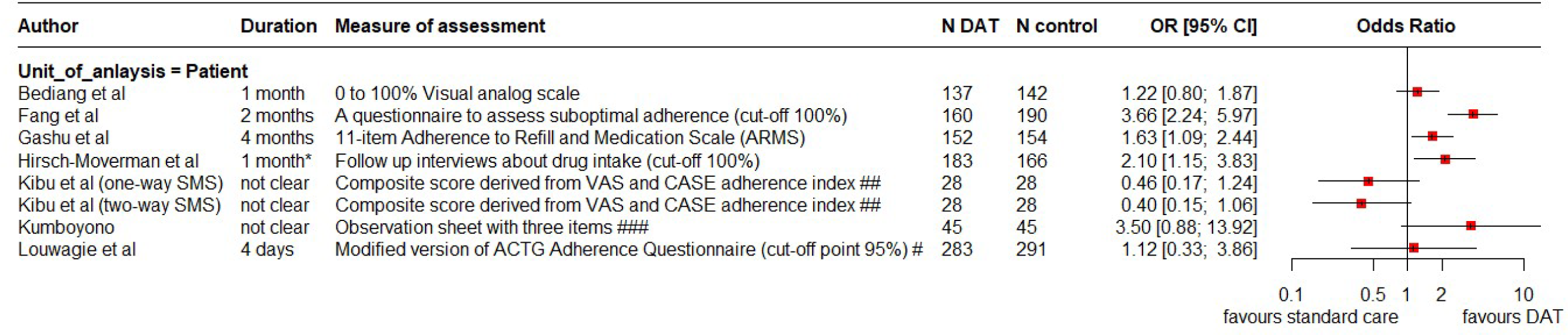
Forest plot showing self-reported adherence to TB treatment in SMS groups versus standard of care. * Adherence was assessed monthly in follow-up visits, # ACTG AIDS Clinical Trials Group. An adherence index was calculated using the 4-day recall formula [doses taken/ doses prescribed], ## VAS Visual Analogue Scale; (CASE) Center for Adherence Support Evaluation, ### observation sheet including three items: the number of drugs consumed; the type of drugs; the time of drug consumption. Patients who satisfy all three subjects are considered adherent

Of 17 studies reporting VOT interventions, 12 (71%) explicitly assessed treatment adherence, based on the videos reviewed, and usually described by the ratio of observed to prescribed doses. Some studies reported the proportion of doses, while others used categorical cut-offs of 80% or 95% to define adherence. Most associated VOT use with improved adherence. (Fig 8)

**Fig 8.**
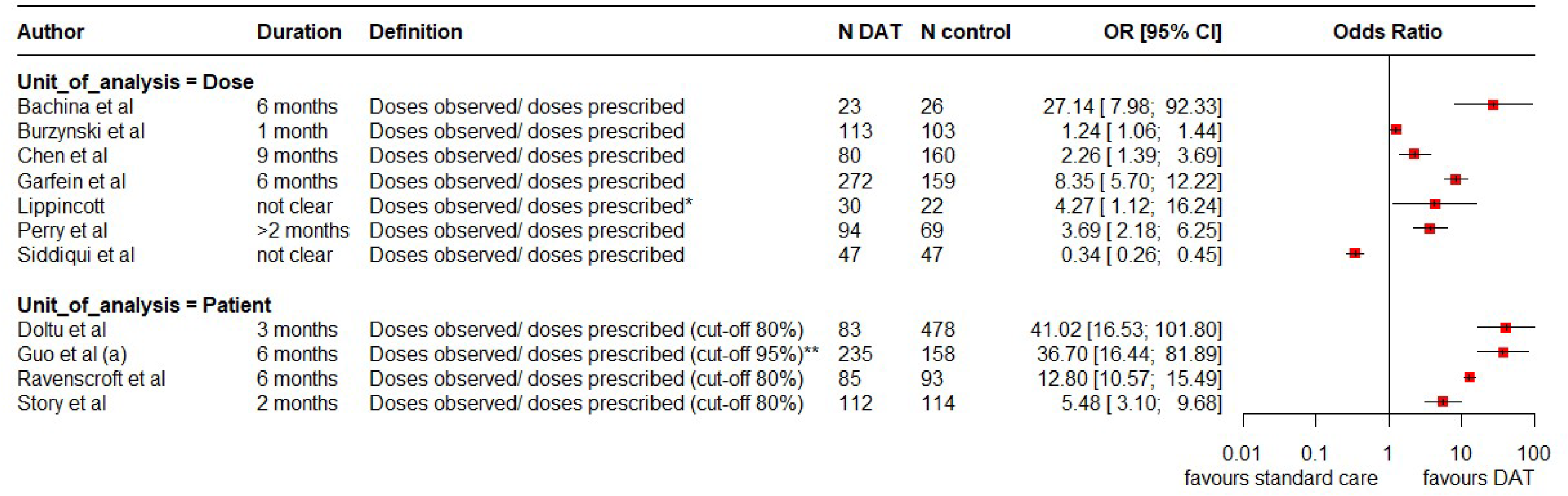
Forest plot showing comparing dose observation in VOT groups versus standard of care. * Prescribed doses that could not be confirmed as ingested were recorded to be ‘missed’ or ‘self-administered’ based on chart documentation, ** The numerator includes either directly observed or video-observed doses, excluding those obtained by self-report. The denominator includes the self-administered medications; it does not include periods when the treatment was suspended based on medical advice because of side effects.

Similarly, among 18 studies involving digital pillboxes, 13 (72%) assessed adherence of which 10 (55%) studies used the pillbox data (Fig 9). Some studies also used complementary measures such as pill counts, self report by persons on treatment, or urine tests.[17,23,31] Most suggested that the use of the pillboxes was associated with improved adherence.

**Fig 9.**
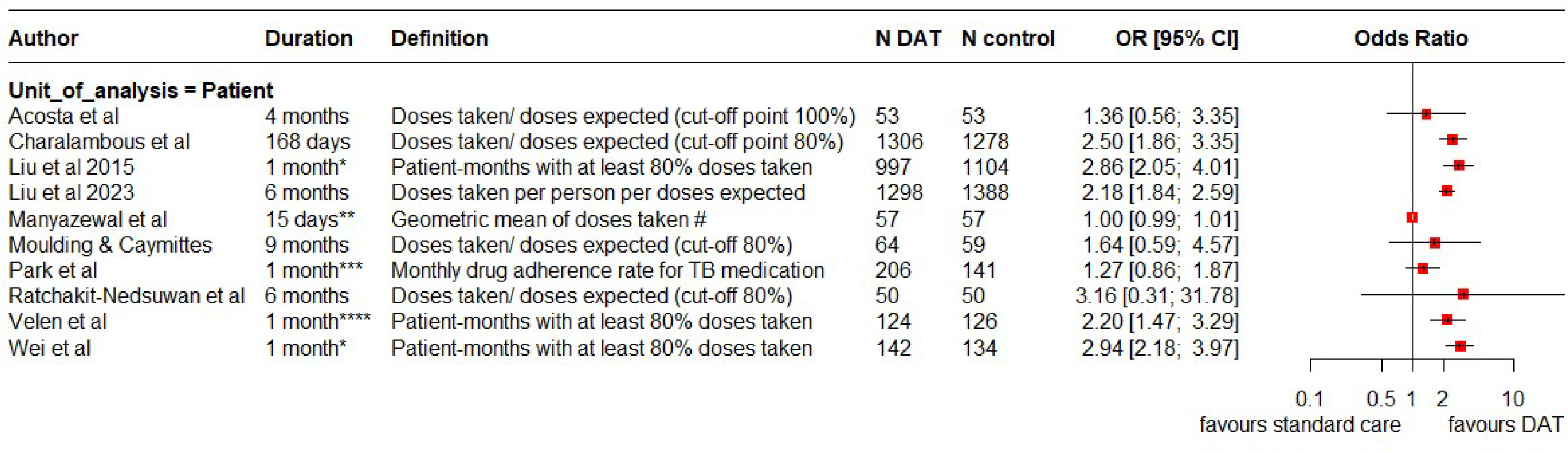
Forest plot showing adherence to TB medication dosing from digital pillbox data in digital pillbox group versus standard of care. * The study provided the percentage of patient-months on TB treatment where at least 20% of doses were missed. Adherence was measured for 6 months, ** Four 15-day appointments (intensive phase), *** The study provided the mean monthly drug adherence. Adherence was measured monthly for 6 months, **** Adherence was measured monthly for 6 months, # individual-level % adherence averaged over the 2-month intensive phase measured

Feature-phone based interventions were generally associated with improved adherence.[33,34] For interventions that used smartphone technology, results were inconsistent. While one study[35] suggested that the technology was associated with an improvement in treatment adherence of persons with multi-drug resistant TB, the two other studies found conflicting results among persons with drug-sensitive TB. [36,37]

### Patient-reported outcomes

#### Satisfaction

Satisfaction of people with TB with their care was assessed in 2 SMS studies.[38,39] One study assessed satisfaction at the end of the sixth month on treatment with a Likert scale.[38] In this study, satisfaction appeared high and similar between SMS and standard of care groups based on two general questions. Another SMS study [39] assessed satisfaction with the provider–patient relationship using a more detailed 7-item questionnaire; satisfaction was significantly higher in the intervention group than in the control group. (S20 Table)

Three VOT studies [19,20,40] used surveys to measure satisfaction among persons with TB; all reported greater satisfaction among participants in the VOT groups.

Only one study [18] reported satisfaction of persons with TB with the digital pillbox, using the Treatment Satisfaction Questionnaire for Medication (TSQM). It includes 14 questions across four domains: Effectiveness, Convenience, Side Effects, and Global Satisfaction. Satisfaction was significantly greater in the DAT group than in the control group in all domains assessed. (Table 17)

#### Health related quality of life

Four studies (one SMS, one VOT, and two digital pillbox studies) assessed health related quality of life (HRQOL) in persons with TB disease [16,19,34,41] using DATs, while one SMS study evaluated HRQOL in persons with TB infection [26]. All five studies used versions of the EuroQol questionnaires and found no differences between participants in the DAT and standard care groups.

#### Stigma

One study measured TB-related stigma from the patient and family perspectives using the Van Rie stigma scale. [34] It did not identify any significant difference in stigma reported by persons with TB and their families supported by a feature-phone intervention vs. those who received standard care.

### Special populations

Other prespecified stratified analyses involving children, persons living with HIV, those with drug resistant TB disease, and those with extrapulmonary TB disease were not conducted as no sufficiently homogeneous studies were found for each subgroup.

## Discussion

Our systematic review aimed to assess the impact of digital technologies on clinical outcomes, adherence, and patient-reported outcomes among persons treated for TB disease and infection. We included randomized and quasi-experimental studies, as well as cohort studies, from diverse country settings.

Some digital technologies, notably those involving video observation or other smartphone technologies, were associated with improved treatment success and reduced losses to follow-up. They may also enhance adverse event reporting. These findings are concordant with previous reviews [5,8,42,43]. Similarly, VOT was associated with improved adherence to expected dosing. However, VOT and smartphone intervention studies were overwhelmingly conducted in high-income and upper-middle-income countries. Hence the generalizability of these findings to low-income and lower-middle-income country settings, which experience most of the world’s TB incidence, may be limited. Low-income and lower-middle-income countries may face greater challenges in mobile coverage and accessibility, and in health system capacity to implement DATs.

Indeed, SMS-based interventions and medication sleeves with phone calls (99DOTS), which were largely implemented in LMIC settings, were not consistently associated with improved adherence or clinical outcomes. In fact, 99DOTs may have been associated with an increased frequency of death, raising the possibility of gaps in patient supervision and support.[44] However, this result should be interpreted with caution as the effect of confounders was not adjusted for. Clustering was also not considered in one of the 99DOTS studies. It is also possible that deaths appeared more frequent if there were fewer losses to follow-up, as the latter are a key competing risk.

Digital pillbox-based interventions, which were also mostly implemented in LMIC settings, also had variable associations with clinical outcomes. While digital pillboxes were associated with increased medication adherence, these findings were often reported from studies that compared adherence data from a “silent” pillbox (i.e., without reminders) in the control arm to a pillbox that provided audiovisual reminders in the intervention arm. This approach may increase risk of detection bias, as people receiving audiovisual reminders may be more inclined to keep medication blister packs within the device, thereby increasing engagement with the DAT. Earlier studies and a forthcoming systematic review have highlighted limits to the accuracy of dose reports with 99DOTS and reveal considerable variability in the accuracy of dose reports from digital pillboxes. [45–47]

The increased reporting of adverse events with the VOT and smartphone platforms is likely a positive consequence of more consistent opportunities for contact with providers. Reporting of more adverse events by persons treated using VOT may indicate that more medication is taken. Reporting of adverse events is important to manage potential complications, and to address concerns that may affect adherence to treatment.[20]

Unfortunately, no study evaluated deaths post-treatment, and most did not report on recurrent TB disease or emergence of drug resistance at any time.

Importantly, many of the VOT and digital pillbox studies also included co-interventions to enhance patient support, beyond dose monitoring. Examples include medication reminders,[19,22,25,28,40,48–51] encouraging adverse event reporting [19,20,22,24,40,48] and/or escalation via SMS, phone calls, or home visits if expected video observation was missed. Some VOT interventions were coupled with motivational SMS, reimbursement of mobile data costs, payment for visit attendance, or nutritional allowances. [19,20,29,48,52] Similarly, for interventions that involved smartphone technologies, most studies described various combinations of associated measures e.g., medication reminders, adverse event reporting, instant messaging, chat groups with providers and/or other patients. [37,53,54]

The diversity of comparison strategies reported as the “standard of care” further complicates interpretation and inference. While most studies indicated daily provider-administered DOT (facility-or field-based) as the comparator, others used family DOT or self-administered treatment, a combination of approaches, or simply did not specify.

Strengths of this review include a comprehensive search, which ultimately yielded a large number of included studies (n = 70), with many participants (n = 58,797) in diverse settings, permitting more detailed, stratified analyses. We also assessed the potential association of DATs with a variety of outcomes.

However, our review has several limitations. Most studies reporting VOT or other smartphone technology-based interventions were conducted in high- or upper-middle income countries. This substantially limits generalizability of findings regarding these two technologies to lower-income country settings where TB burden is highest, yet where these technologies and the necessary technical and human resource infrastructure may be least accessible. Notably, however, our review does report robust findings from studies in LMICs: the DAT types implemented in these settings (e.g., SMS, 99DOTS, digital pillboxes) had variable or non-significant impacts on treatment outcomes, which has important policy implications.

There was also substantial heterogeneity in study findings, which is not surprising given the diversity of study settings, methodologies, technologies and comparators considered. This limited our ability to formally pool and meta-analyze data, and made some quantitative findings less robust.

We could not assess treatment success and treatment completion as separate outcomes: some studies used the two terms interchangeably. While the treatment success category includes both persons with microbiologic cure and those who completed treatment without known microbiologic failure, most studies did not report microbiologic cure.

In this meta-analysis, we estimated odds ratios for some short-term clinical outcomes from crude event numbers reported in cluster randomized trials, as no adjusted estimates were provided. This may have resulted in artificially narrow confidence intervals when clustering was ignored.

We focused on evidence from RCTs, quasi-experimental studies and cohort studies. For this reason, cross-sectional studies which assessed patient-reported outcomes—often without a comparison group—were excluded. More generally, there was very limited evidence around the use of DATs in persons with drug-resistant TB, in children, or in persons living with HIV.

## Conclusion

While the evidence base remains incomplete, and generalizability of studies often limited, this systematic review nonetheless synthesizes relevant information and identifies further evidence gaps. It provides insights into how some DATs can be successfully used to support curative and preventive TB treatment. VOT and smartphone interventions were associated with increased treatment success in high-income settings and should strongly be considered together with digital pillboxes as alternatives to DOT when appropriate. SMS, 99DOTS, and other feature-phone based interventions did not improve treatment success or had highly variable findings, with most studies conducted in LMIC settings. This finding has important policy implications and raises questions regarding investments in the DATs as currently designed and implemented. With the rapid growth of technology and mobile usage worldwide, programs, patients, and providers will increasingly look to digital technologies for support. The combination and fine-tuning of different support interventions, with a more personalized delivery approach, is promising. However, there remain substantial knowledge gaps with respect to key populations and to longer-term health outcomes. Beyond issues of generalizability to settings and populations which bear the highest TB burdens, there also remain important questions related to implementation and logistics. Further research is urgently needed to rethink approaches to DATs—and to inform non-DAT-based approaches to improving TB treatment outcomes in LMICs.

## Data Availability

Most relevant information is within the Supporting Information files. However, all raw data and instruments will be posted publicly on the McGill International TB Centre website https://www.mcgill.ca/tb/ following acceptance for publication.

